# Automated Detection Model in Classification B-Lymphoblast Cell from Normal B-Lymphoid Precursors in Blood Smear Microscopic Images Based on the Majority Voting Technique

**DOI:** 10.1101/2021.07.10.21260312

**Authors:** Mustafa Ghaderzadeh, Azamossadat Hosseini, Farkhondeh Asadi, Hassan Abolghasemi, Arash Roshanpour

## Abstract

**Introduction:** Acute Lymphoblastic Leukemia (ALL) is a deadly white blood cell disease that affects the human bone marrow. Detection of ALL, the most common type of leukemia, has been always riddled with complexity and difficulty in its early stages. Peripheral blood examination as a common method at the beginning of the ALL diagnosis process is a time-consuming, tedious process and greatly depends on the experts’ experience, keeping up with the advances in artificial intelligence in the diagnosis process. Keeping up with the growth and development of artificial intelligence algorithms a model was developed to classify B-ALL lymphoblast cells from lymphocytes.

**Materials and Methods:** A Fast, efficient and comprehensive model based on Deep Learning (DL) was proposed by implementing eight well-known Convolutional Neural Network (CNN) models for feature extraction on all images and evaluating in classifying B-ALL lymphoblast and Normal. After evaluating their performance, four best-performing CNN models were selected to compose an ensemble classifier, by combining the model performance of each classifier.

**Results:** Due to the close similarity of the nuclei of cancerous and normal blood B-ALL cells, the state-of-the-art CNN models alone did not achieve acceptable performance in diagnosing these two classes and their sensitivity was low. The proposed classification model Based on the majority voting technique was adopted to combine the CNN models. The sensitivity of 99.4, the specificity of 96.7, AUC of 98.3, and accuracy of 98.5 were obtained for the proposed model.

**Conclusion:** To classify blood cancerous cells from normal cells, the proposed method can achieve high accuracy without the intervention of the operator in cell feature determination. Thus, the DL-based model can be recommended as an extraordinary tool for the analysis of blood samples in digital laboratory equipment to assist laboratory specialists.

## 1. Introduction

Leukemia is one of the most common blood cancers that affected 174,000 people in 2018 in the United States alone. Research shows that about 6,000 cases of leukemia are diagnosed each year, of which acute lymphoblastic leukemia (ALL) is the second most common type in adults and the most common type of malignancy in children, accounting for about one-third of all childhood cancers. Based on the most recent World Health Organization (WHO) classification, the purely leukemic presentation, B-lineage ALL (85 %), is the most common type of lymphoid cancers^1^. Acute lymphoblastic leukemia type B (B-ALL) is a neoplastic disorder with high mortality rates due to immature lymphocyte B-cell proliferation. The identification of the signs and symptoms of childhood cancer is very challenging because it is not the first diagnosis to be made for non-specific complaints, leading to potential uncertainty in the diagnosis. Moreover, childhood cancer can mimic other common childhood diseases and even normal growth processes^2^.

B-ALL can be diagnosed via different techniques as announced by the WHO, but this diagnosis is usually performed manually in hospitals by determining the number of leukocytes in peripheral blood stains by immunohistochemical staining. The laboratory staff performs the staining process manually, so this method is highly time-consuming and error-prone. Primary prevention measures are not effective in averting the development of B-ALL in this age group, and secondary prevention, i.e., early diagnosis, is therefore essential. In the specific case of ALL, early diagnosis and treatment increase the chances of cure. To minimize human intervention and overcome the early detection limitations of ALL, the use of computerized tools can effectively minimize the destructive impacts of cancerous tissues on patients.

In the last two decades, many studies have been conducted using machine learning (ML) methods and computer-aided diagnostic methods for the analysis of laboratory images to overcome the limitations of the late diagnosis of leukemia and determine its subtypes. These studies analyzed leukocyte nuclei in blood smear samples to diagnose and differentiate B-ALL from normal leukocytes.

Recently, numerous computer-based methods have been employed to improve the efficiency of medical imaging techniques. One such method is the application of ML algorithms which has achieved remarkable success in medical imaging. Among different types of ML methods, deep learning (DL) models have attained a high precision in machine vision tasks in leukemia. Convolutional neural networks (CNNs) have high potential in feature extraction and analysis. Based on the current trend, DL models have obtained high accuracy scores in computer visual tasks, and this has motivated research into the use, adaptation, and application of such models in medical image analysis^3^. Due to the limitations of diagnostic tests, numerous ML techniques have been adopted to improve the precision of diagnostic methods. Table (1) lists some relevant studies.

**Table 1:**
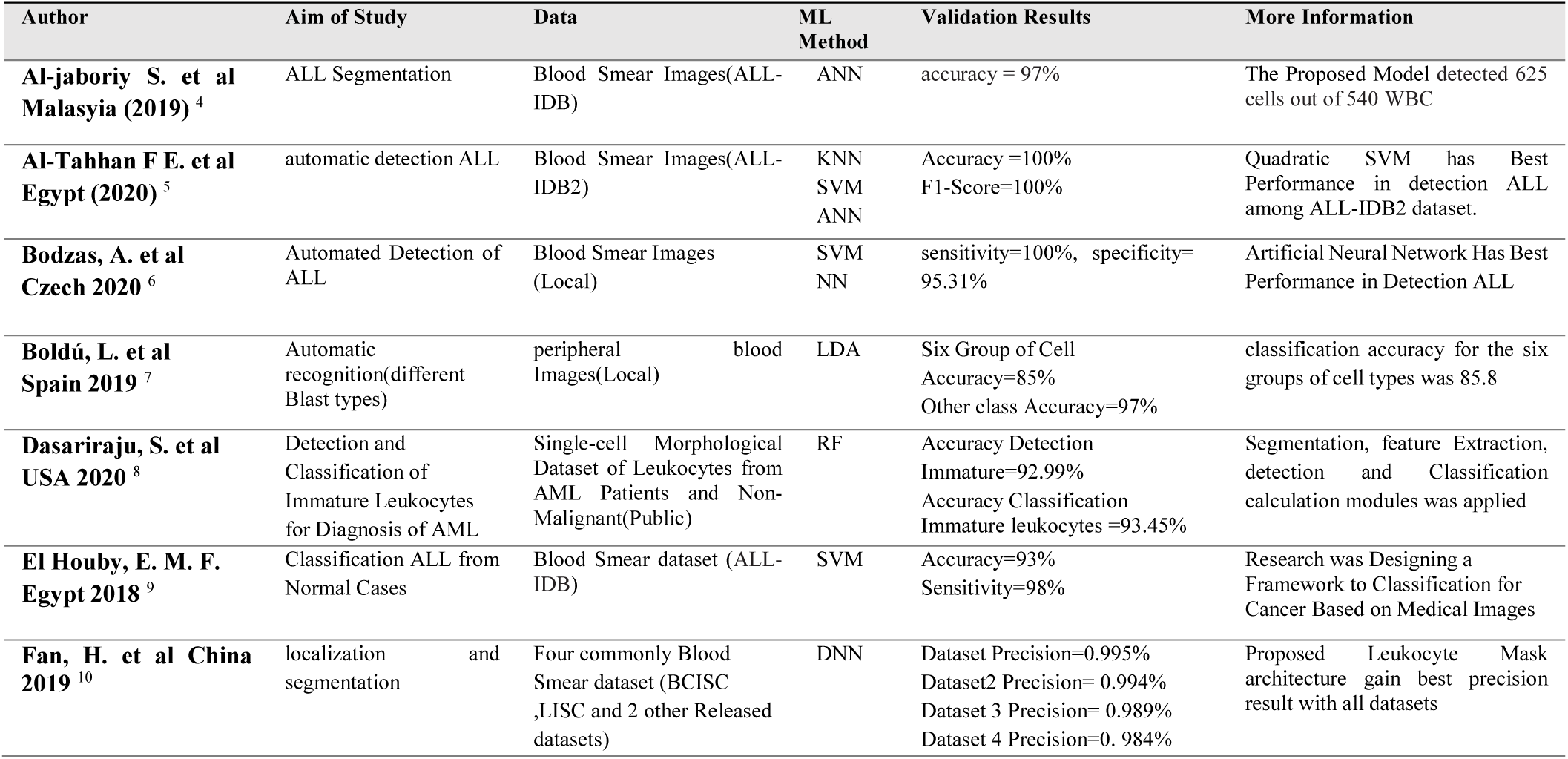
Characteristics of Studies using ML Algorithms in the Detection and Classification of Blood Smear Images. The reason for choosing DL as the selection algorithm of this research is that feature extraction is performed automatically, and the researcher plays no role in arbitrary feature selection. In these methods, building blocks of convolution neural network, including convolution and pooling layers, processes the values corresponding to the pixels; in this way, features are extracted automatically. Then, the features are classified by feeding the features to a layer containing one or more classifiers. For the diagnosis of blood diseases, researchers have considered the use of DL and its main algorithm, CNN. In this regard, by adopting a hybrid model, B lymphoblasts are detected, differentiated from normal B lymphocyte precursors, and classified. Herein, we introduce a novel nucleus detection method based on a CNN pre-trained model in which, by using four state-of-the-art CNNs, a model is proposed for the classification of B-ALL from normal lymphocytes.

## 2. Materials and Methods

Based on recent research, DL algorithms have brought positive potential into image processing as an optimal solution.

### 2.1 Dataset

The C-NMC dataset^11–14^ comprised 12528 lymphocyte nucleus images, of which 8491 belonged to B-ALL lymphoblast and 4037 to normal B-lymphoid cases. The dataset cell nuclei were segmented from the microscopic images in the real world because these contain some staining noise and illumination error, although an expert via an in-house method of stain color normalization has largely fixed these errors. An expert oncologist marked the ground truth of the dataset images. Figure (1) illustrates samples of B-ALL and healthy cell nuclei.

**Figure 1.**
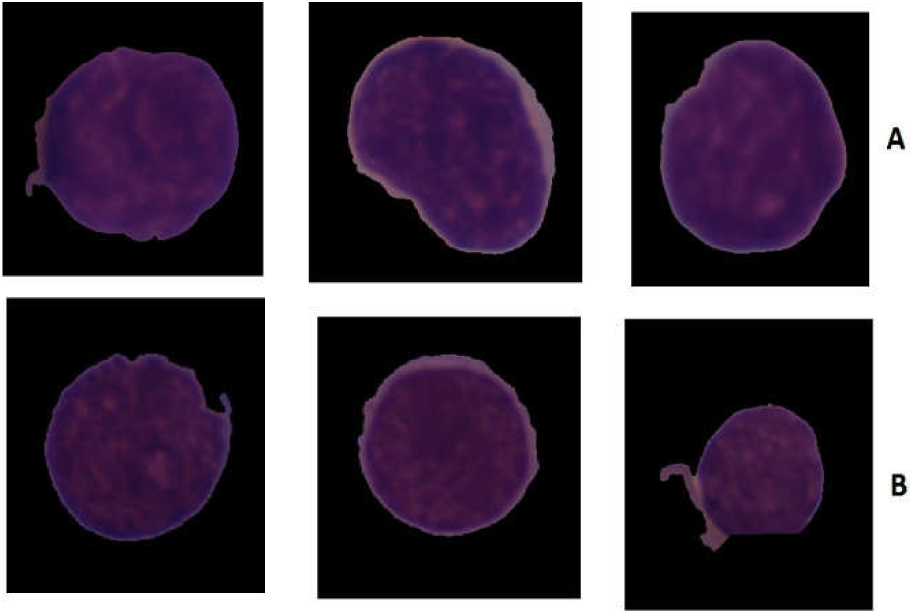
A) B-ALL lymphoblast nuclei, B) Normal cell lymphocyte nuclei

Motivated by the success of CNNs in machine vision, this study designed a detection model for the classification of cell nucleus images belonging to B-ALL lymphoblast from B-ALL lymphocyte cases and implemented it by presenting a CNN based on different architectures feature extractors.

### 2.2. Data Preparation and Preprocessing

According to the scientific texts, Data standardization and normalization as the first steps of preprocessing to maintain image integrity plays a key role in image analysis and classification ^15,16^. To this end, first, the pixel-level global mean and standard deviation (STD) were calculated for all images, and then the data were normalized using Equation (1), where 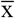 denotes the global mean X of the image set, σ is the STD, and ε = 1e -10 indicates the differential value to prevent the denominator from becoming zero.

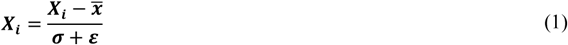

After normalization, to standardize the image for achieving a uniform ratio for the input of the deep neural network, the pixel value of each image is mapped to [0,255] and then converted to the [0,1] interval, so that the image becomes a standard image since the CNN method relies on large amounts of data to improve its efficiency and prevent model overfitting ^17,18^. Given that in this study, we were dealing with the WBC core. Hidden features in the WBC core such as the density of the core, the smoothness and serration of the core wall, and so on. After normalization and standardization of images, the core of these images was enlarged by cutting the edges of the image so that image processing algorithms could analyze the characteristics of different classes more easily by analyzing the nucleus of lymphocytes. Figure 2 shows the two operations of cutting the edge and enlarging the core. Data augmentation was performed for the training dataset by 16 techniques for each image.

**Figure 2:**
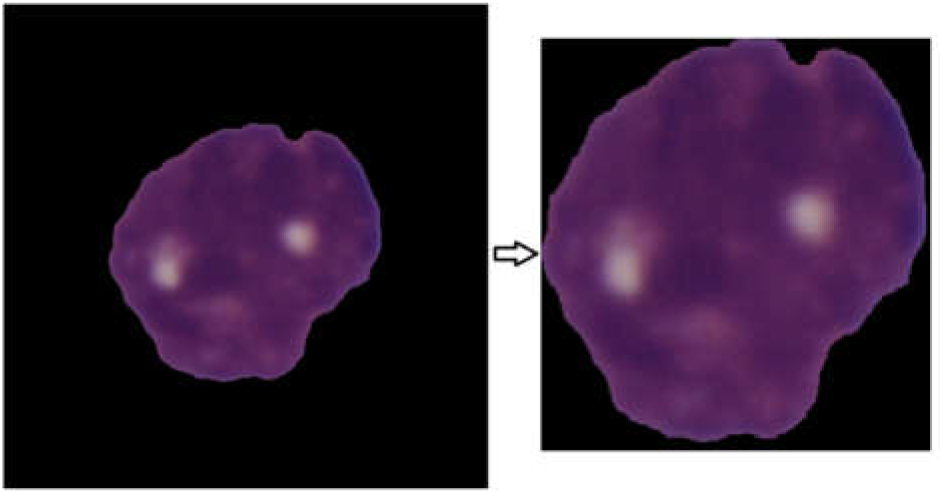
Image edge cutting to enlargement the morphological features

Whereas in this study, we are dealing with the nucleus of white blood cells. The hidden features of the WBC core include the density of chromatin, open chromatic of nuclei and Nucleolus, and other features.

All the images in these collections were shuffled so that, during the training process, the network would not see only specific categories of data, and each batch of images would contain different labels belonging to B-ALL and non-B-ALL categories. The size of the input image was changed to 300×300×3, but this method can be used on images with any size. The dataset was divided into training, validation, and test sets, according to the default hypothesis of ALL challenge dataset of ISBI 2019, whereby 10,661 of the data (ALL (cancer): 7272, normal: 3389, including 10% of the training dataset for validation) were allocated to the training set, and the remaining 1847 to the test set.

### 2.3 Deep Learning Algorithms

For medical image segmentation, numerous CNN-based methods are widely used benefitting from its powerful feature learning and representation ability. Of these methods, the convolutional Neural network (CNN) has demonstrated state-of-the-art performance in cells and organ segmentation problems^19–21^. CNN is a multi-layer network composed of overlapping convolutional layers (for feature extraction) and down-sampling layers (for feature processing). Figure 3 shows the structure of a typical convolutional neural network. CNN can automatically extract features from images, so it has become a research hotspot. CNN is a perceptron-based mode. CNNs can automatically extract features from images, and thus have become a research hot topic. CNNs are perceptron-based models^22,23^.

**Figure 3:**
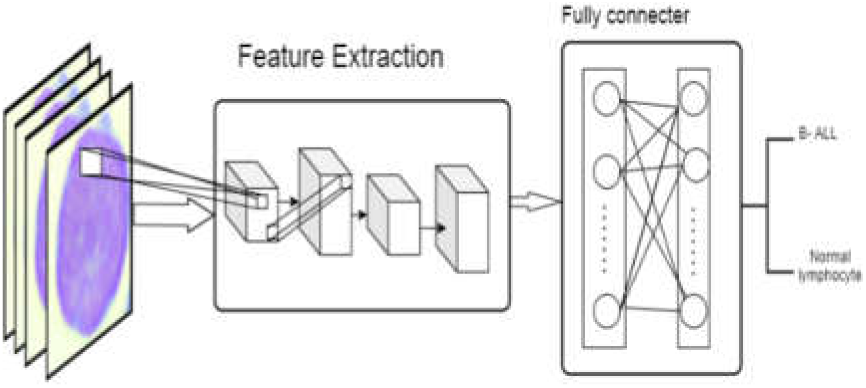
An ordinary convolutional neural network structure

Pre-trained models who have been trained in large image collections have achieved extraordinary results in their classic image issues and are therefore referred to as the State of the art. Many studies have used these top-notch models because their ability to extract image features has outperformed other models. Among the many publicly available deep learning pre-training models, Alexnet^24^, ResNet^25^(ResNet50, ResNet101 Inception-V3^25^, Inception-ResNet-V2^26^, SqueezeNet^27^, and MobileNet-V2^28^ Used and compared, because these network models show higher performance in terms of accuracy compared to any other networks with similar prediction times. Those well-known CNN models were pre-trained with the ImageNet database.

Densenet-121 was presented by Huang to make DL programmable by eliminating the problem of missing gradients, increasing feature reuse, and improving parameter efficiency. It has achieved advanced performance in several computer vision tasks. In addition, DenseNet-121 has successfully used medical images to predict diseases^29^. Moreover, Inception-ResNet-v2 is a combination of two recent networks, residual connections and a recent version of the Inception architecture. The residual model is well-known for training very deep architectures. The hybrid InceptionResNet-v2 network utilizes residual connections with good efficiency^25,30,31^. The last pre-trained network, Xception, is a CNN based on depth wise separable convolution layers. These well-known architectures were chosen due to their success in processing medical images. The process commenced with two convolution layers, followed by depth wise separable convolution layers, four convolution layers, and a fully-connected layer^32^.

### 2.4 Ensemble Learning

Ensemble methods are algorithms that take the advantage of an ensemble of classifiers to predict labels of data. The weighted majority voting algorithm was introduced by Littlestone and Warmuth in 199432. Its idea depends on the final decision result by taking the weighted majority votes of the other algorithms. They have proved that the ensemble methods are robust algorithms with respect to the error and can significantly improve the generalization ability of the learning system. The algorithm forms its prediction by comparing the total weights made for each class and predicts the larger total. In this way, the classification result is voted to obtain the final classification result. The voting method is divided into absolute and relative majority voting methods. Absolute majority voting method, that is, where more than half of individual learners output the same classification result, the result is the final classification of integrated learning output.^33^

### 2.5 Performance evaluation

To assess the performance of the CNNs models, four performance indices were calculated. The model’s performance was determined by using the confusion matrix^34^. Here, sensitivity was defined as the ratio of B-ALL cases correctly detected by the model to all the actual B-ALL lymphoblast cases. Specificity is the ratio of the normal lymphocyte cases correctly detected by the model to all the actual non-B-ALL (normal) cases. Moreover, accuracy was defined as the rate of all the B-All and lymphocyte cases accurately classified. We used traditional measures to evaluate the performance of the proposed model based on the confusion matrix. According to the confusion matrix, the specificity and sensitivity to measure and analyze the performance of the network can be calculated^35^. Specificity is the classifier’s ability to correctly identify cases without the disease (true negative rate), while sensitivity is the classifier’s ability to correctly classify all the cases with the disease (true positive). The formulas for the evaluation criteria are given in Equations (2-4):

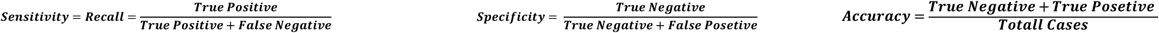

## 3. Results

By implementation, seven models with learning data sets and then testing these models using a test dataset. It was concluded that different states of the art shown different performances. With respect to the similar features of the two data classes, the models did not reach an acceptable performance. The evaluation metric for the implemented network based on the confusion matrix is given in Table (2).

**Table 2:**
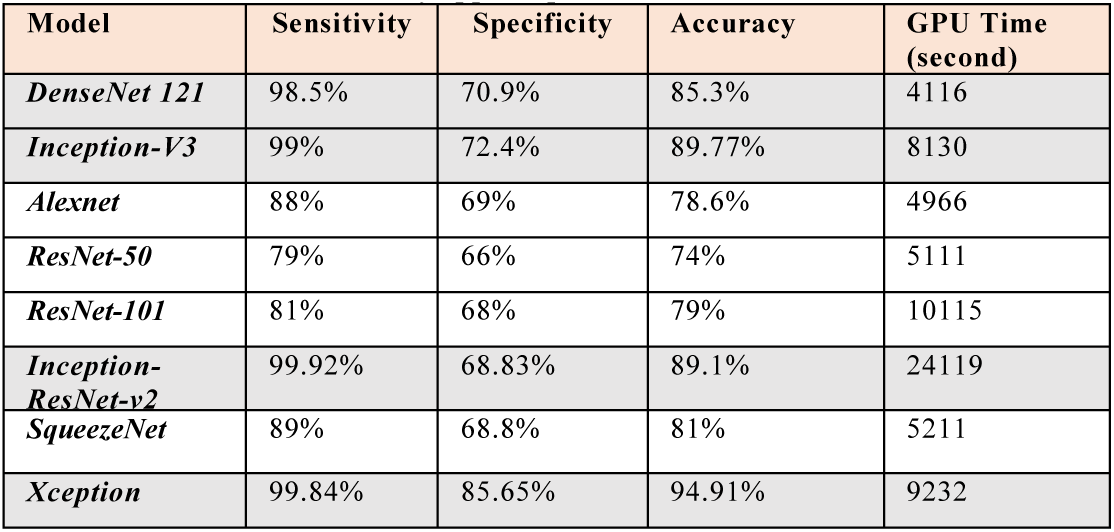
Evaluation metrics of applied pre-Trained Models

| Model | Sensitivity | Specificity | Accuracy | GPU Time (second) |
| --- | --- | --- | --- | --- |
| DenseNet 121 | 98.5% | 70.9% | 85.3% | 4116 |
| Inception-V3 | 99% | 72.4% | 89.77% | 8130 |
| Alexnet | 88% | 69% | 78.6% | 4966 |
| ResNet-50 | 79% | 66% | 74% | 5111 |
| ResNet-101 | 81% | 68% | 79% | 10115 |
| Inception-ResNet-v2 | 99.92% | 68.83% | 89.1% | 24119 |
| SqueezeNet | 89% | 68.8% | 81% | 5211 |
| Xception | 99.84% | 85.65% | 94.91% | 9232 |

Training these seven well-known models with training data sets and then validating them, four models achieved higher performance. These best four models (DenseNet121, Inception V3, Inception-ResNet-v2, and Xception) among eight models were selected by evaluating the performance of each model in terms of accuracy and calculation time.

### 3.1 Ensemble deep learning model for classification B-ALL form B-lymphoblast (Normal)

In this study, by examining the performance of the models used, the researchers sought a way to improve the results of these models. The majority-voting group was used as the final decision for this model. Present model scheme first calculates the total number of votes received by each base classifier, than the majority of votes is calculated by the classification of both class. Algorithm 1 presents the proposed model in detail. Let *L= {*DenseNet121, Inception V3, Inception-ResNet-v2 and Xception} be the set of pre-trained models. Four used selection model is fine tuned with the images from training datasets (*X; Y*); where *X* the set of *N* images, each of size, 300 × 300, and *Y* contain the images labels, *Y is a collection of two class including Lymphoblast B-ALL and Normal B-ALL*. After dividing the training dataset into batches size n=256.

**Algorithm 1:**
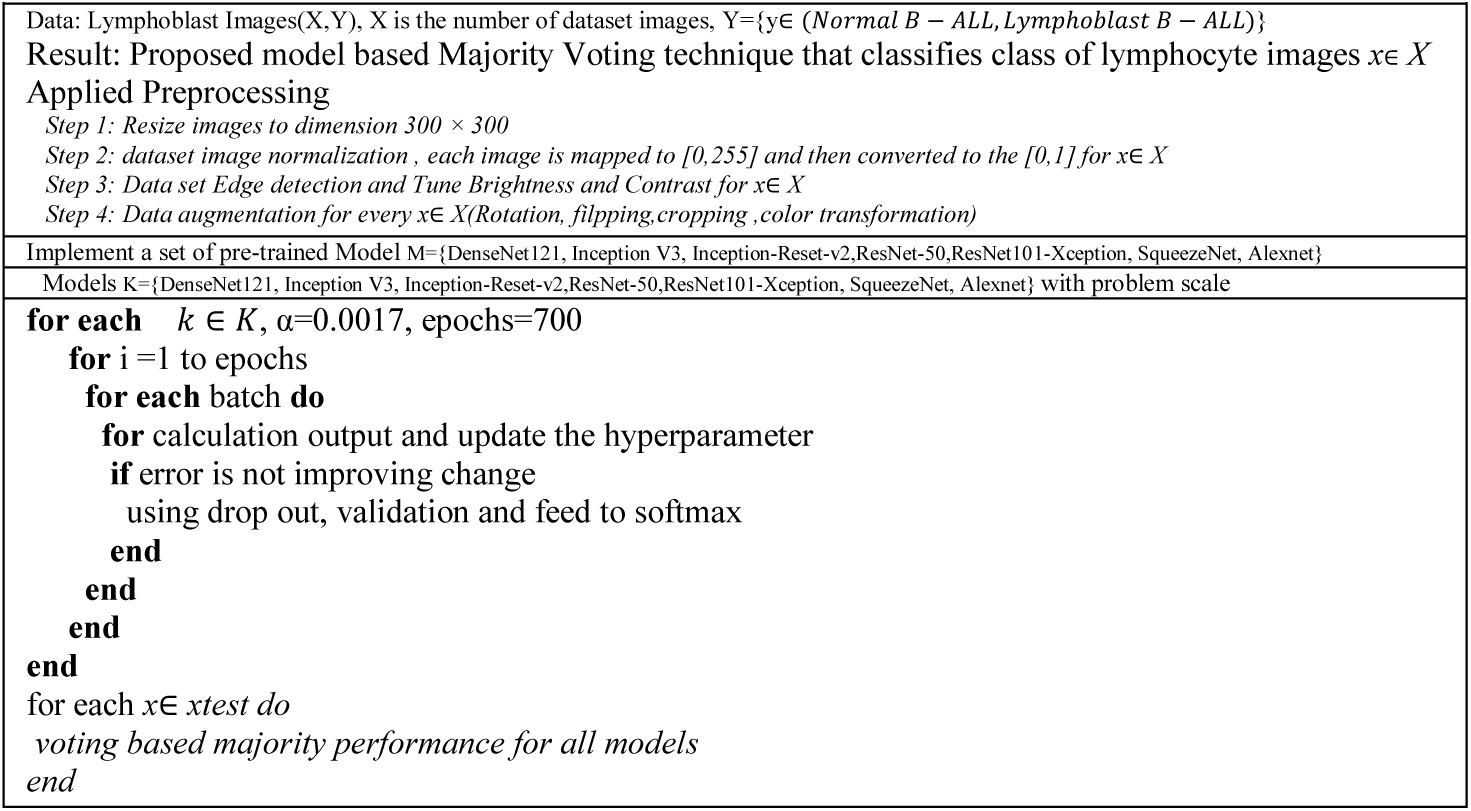
Proposed Ensemble Algorithm

In this method, the number of base classifiers should always be odd. In the case of an equal vote, the mode function applies is applied.^36^ In this section, the proposed B-ALL disease detection and classification model is presented. The proposed model is displayed in Figure 4. It is an ensemble framework of four models.

**Figure 4:**
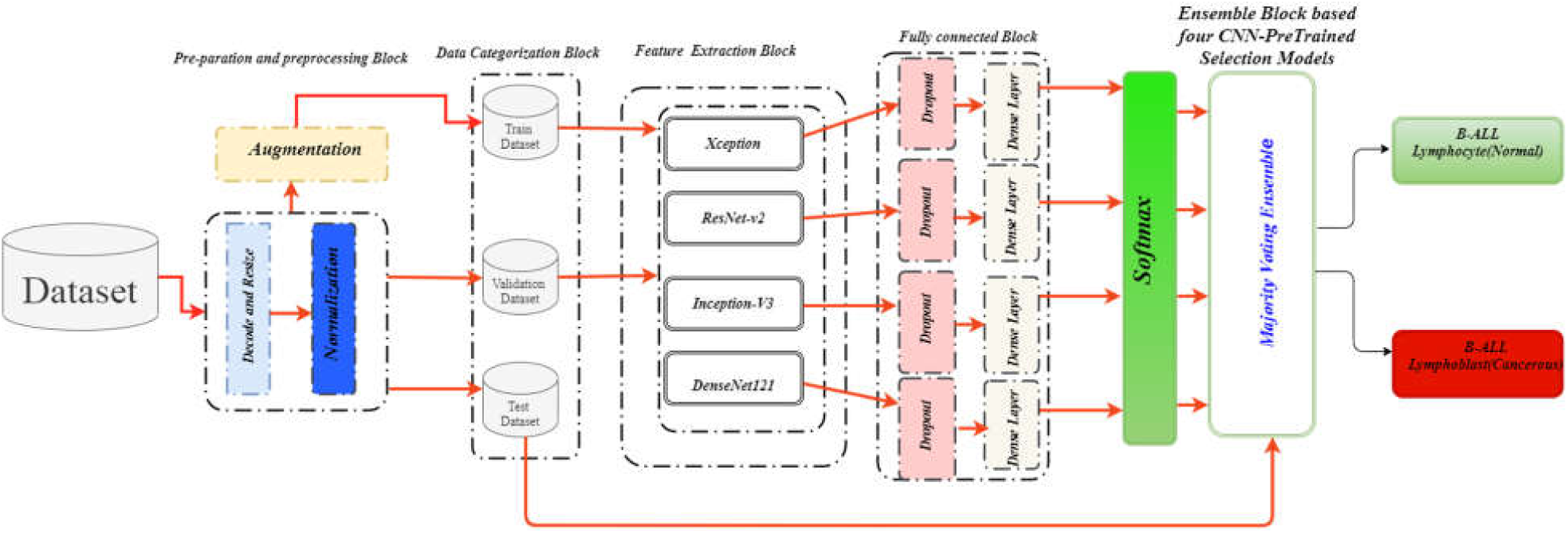
Proposed model for the classification of B-ALL lymphoblast from B-ALL lymphocytes

These four hybrid models are in parallel and combine module the output by the ensemble technique to improve the classification confidence and accuracy. Based on the confusion matrix in Figure 5, specificity and sensitivity are presented in Table 3 to assess the performance of the proposed ensemble model. The accuracy of the proposed model is 98.5% and the f1–score is 98.3%.The performance of the classification results of the proposed ensemble model and the previous state-of-the-art models is compared in Table (3). Evidently, the proposed ensemble model has achieved a promising performance that is superior to the previous models. The success of the proposed model with such a small data set is attributed to the use of class weights in the training process.

**Table 3:** Performance comparison of different models for ALL the detection

| Researcher, Year | Sensitivity | Specificity | Accuracy |
| --- | --- | --- | --- |
| Mohapatra S et al. (2013) | 94.93 | 95.00 | 94.73 |
| Singhal et al. (2014) | 80.43 | 95.08 | 89.72 |
| Shafique S et al. (2018) | 96.74% | 99.03 | 96.06% |
| Yu et al. (2017) | 88.5% | 77.85 | 98.1 |
| Pansombut T et al.(2019) | 81.56 | Not Mentioned | 81.74 |
| Hosseinzadeh S. et al(2019) | 95.17 | 98.58 | 96.17 |
| **Proposed Model** | **99.4** | **96.7%** | **98.5%** |

**Figure 5:**
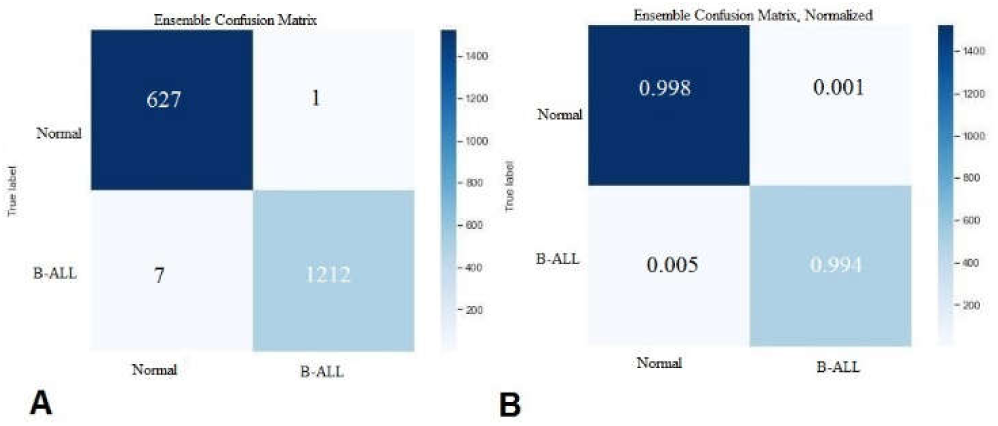
A. Confusion matrix, and B. Normalized confusion matrix of model performance

## 4. Discussion and Conclusion

The analysis of peripheral blood smear images plays a vital role in the diagnosis of various types of leukemia, anemia, and malaria. Unusual alterations in the color, shape, and size of blood cells indicate an abnormal condition. The results of this assessment, which is usually performed manually, depend on the skill and experience of the technician. Besides, manual microscopic examination is time-consuming and yields poor results^37–39^. The proposed method presented a framework for the automated classification of leucocyte cell nuclei from microscopic peripheral blood smear images.

This study introduced a novel combination of image processing methodologies based on state-of-the-art CNNs. The ensemble framework based on majority voting for B-ALL detection using blood smear images achieved high accuracy in the classification of the two classes. Ensemble learning can significantly improve the generalization ability of the learning system, thereby enhancing the performance of the available methods.

## Supporting information

word file

## Data Availability

Data is public and accessible

https://biomedicalimaging.org/2019/challenges/

## 5. Acknowledgments

Not applicable.

## Authors’ contributions

Conceptualization:MG,AH.Programming of the frameworks:MG,AR,FA.Medical Supervisor:DB,HA. Writing,review and editing:MG,AH,FA.

## Availability of data and materials

Data are available and public inhttps://wiki.cancerimagingarchive.net/display/Public/C_NMC_2019+Dataset%3A+ALL+Challenge+dataset+of+ISBI+2019

## Declarations

### Ethics approval and consent to participate

Not applicable.

### Consent for publication

Not applicable.

