## Supplementary material for "Automated Detection Model in Classification B-Lymphoblast Cell from Normal B-Lymphoid Precursors in Blood Smear Microscopic Images Based on the Majority Voting Technique": word file

Mustafa Ghaderzadeh^a^, Azamossadat Hosseini^a*^
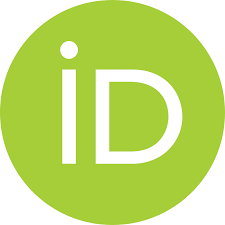
,Farkhondeh Asadi^a^, Davood Bashash^b^,Hassan Abolghasemi^c^, Arash Roshanpour^d^

^a^Department of Health Information Technology and Management, School of Allied Medical Sciences, Shahid Beheshti University of Medical Sciences, Tehran, Iran

^b^Department of Hematology and Blood Banking, School of Allied Medical Sciences, Shahid Beheshti University of Medical Sciences, Tehran

^c^Pediatric Congenital Hematologic Disorders Research Center, Shahid Beheshti University of Medical Sciences, Tehran, Iran

^d^Department of Computer Science, Sama Technical and Vocational Training College, Tehran Branch (Tehran), Islamic Azad University (IAU), Tehran, Iran.

| Table 1: Characteristics of Studies using ML Algorithms in the Detection and Classification of Blood Smear Images | | | | | |
| --- | --- | --- | --- | --- | --- |
| Author | **Aim of Study** | **Data** | **ML Method** | **Validation Results** | **More Information** |
| Al-jaboriy S. et al Malasyia (2019) ^4^ | ALL Segmentation | Blood Smear Images(ALL-IDB) | ANN | accuracy = 97% | The Proposed Model detected 625 cells out of 540 WBC |
| Al-Tahhan F E. et al  Egypt (2020) ^5^ | automatic detection ALL | Blood Smear Images(ALL-IDB2) | KNN  SVM  ANN | Accuracy =100%  F1-Score=100% | Quadratic SVM has Best Performance in detection ALL among ALL-IDB2 dataset. |
| Bodzas, A. et al  Czech 2020 ^6^ | Automated Detection of ALL | Blood Smear Images )Local) | SVM  NN | sensitivity=100%, specificity= 95.31% | Artificial Neural Network Has Best Performance in Detection ALL |
| Boldú, L. et al  Spain 2019 ^7^ | Automatic recognition(different Blast types) | peripheral blood Images(Local) | LDA | Six Group of Cell Accuracy=85%  Other class Accuracy=97% | classification accuracy for the six groups of cell types was 85.8 |
| Dasariraju, S. et al  USA 2020 ^8^ | Detection and Classification of Immature Leukocytes for Diagnosis of AML | Single-cell Morphological Dataset of Leukocytes from AML Patients and Non-Malignant(Public) | RF | Accuracy Detection Immature=92.99%  Accuracy Classification Immature leukocytes =93.45% | Segmentation, feature Extraction, detection and Classification calculation modules was applied |
| El Houby, E. M. F.  Egypt 2018 ^9^ | Classification ALL from Normal Cases | Blood Smear dataset (ALL-IDB) | SVM | Accuracy=93%  Sensitivity=98% | Research was Designing a Framework to Classification for Cancer Based on Medical Images |
| Fan, H. et al China 2019 ^10^ | localization and segmentation | Four commonly Blood Smear dataset (BCISC ,LISC and 2 other Released datasets) | DNN | Dataset Precision=0.995%  Dataset2 Precision= 0.994%  Dataset 3 Precision= 0.989%  Dataset 4 Precision=0. 984% | Proposed Leukocyte Mask architecture gain best precision result with all datasets |

2. **Materials and Methods**

Based on recent research, DL algorithms have brought positive potential into image processing as an optimal solution. The reason for choosing DL as the selection algorithm of this research is that feature extraction is performed automatically, and the researcher plays no role in arbitrary feature selection. In these methods, building blocks of convolution neural network, including convolution and pooling layers, processes the values corresponding to the pixels; in this way, features are extracted automatically. Then, the features are classified by feeding the features to a layer containing one or more classifiers. For the diagnosis of blood diseases, researchers have considered the use of DL and its main algorithm, CNN. In this regard, by adopting a hybrid model, B lymphoblasts are detected, differentiated from normal B lymphocyte precursors, and classified. Herein, we introduce a novel nucleus detection method based on a CNN pre-trained model in which, by using four state-of-the-art CNNs, a model is proposed for the classification of B-ALL from normal lymphocytes.

**2.1. Dataset**

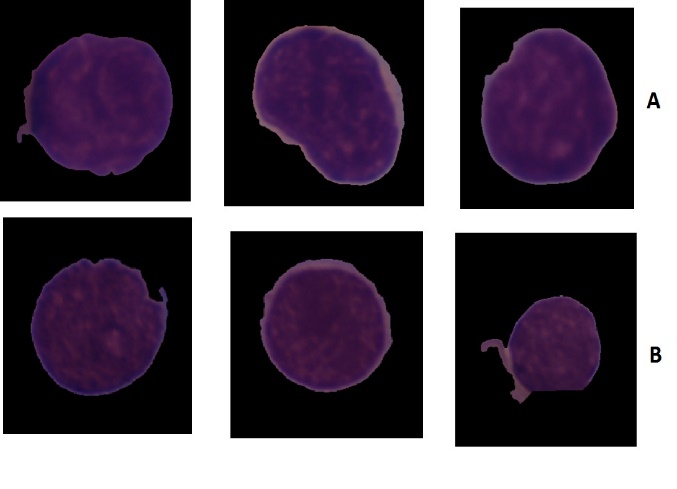
The C-NMC dataset^11–14^ comprised 12528 lymphocyte nucleus images, of which 8491 belonged to B-ALL lymphoblast and 4037 to normal B-lymphoid cases. The dataset cell nuclei were segmented from the microscopic images in the real world because these contain some staining noise and illumination error, although an expert via an in-house method of stain color normalization has largely fixed these errors. An expert oncologist marked the ground truth of the dataset images. Figure (1) illustrates samples of B-ALL and healthy cell nuclei.

Motivated by the success of CNNs in machine vision, this study designed a detection model for the classification of cell nucleus images belonging to B-ALL lymphoblast from B-ALL lymphocyte cases and implemented it by presenting a CNN based on different architectures feature extractors.

Figure 1A) B-ALL lymphoblast nuclei, B) Normal cell lymphocyte nuclei

**2.2. Data Preparation and Preprocessing**

According to the scientific texts, Data standardization and normalization as the first steps of preprocessing to maintain image integrity plays a key role in image analysis and classification ^15,16^. To this end, first, the pixel-level global mean and standard deviation (STD) were calculated for all images, and then the data were normalized using Equation (1), where x̅ denotes the global mean X of the image set, σ is the STD, and ε = 1e -10 indicates the differential value to prevent the denominator from becoming zero.

(1)

$$\boldsymbol{X}_{\boldsymbol{i}}\mathbf{=}\frac{\boldsymbol{X}_{\boldsymbol{i}}\mathbf{-}\bar{\boldsymbol{x}}}{\boldsymbol{\sigma}\mathbf{+}\boldsymbol{\varepsilon}}$$

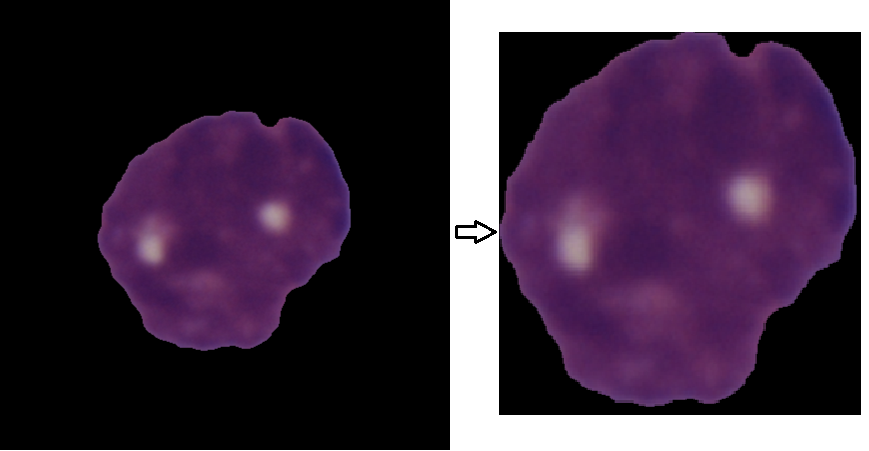
After normalization, to standardize the image for achieving a uniform ratio for the input of the deep neural network, the pixel value of each image is mapped to [0,255] and then converted to the [0,1] interval, so that the image becomes a standard image since the CNN method relies on large amounts of data to improve its efficiency and prevent model overfitting ^17,18^. Given that in this study, we were dealing with the WBC core. Hidden features in the WBC core such as the density of the core, the smoothness and serration of the core wall, and so on. After normalization and standardization of images, the core of these images was enlarged by cutting the edges of the image so that image processing algorithms could analyze the characteristics of different classes more easily by analyzing the nucleus of lymphocytes. Figure 2 shows the two operations of cutting the edge and enlarging the core. Data augmentation was performed for the training dataset by 16 techniques for each image.

Figure 2:Image edge cutting to enlargement the morphological features

Whereas in this study, we are dealing with the nucleus of white blood cells. The hidden features of the WBC core include the density of chromatin, open chromatic of nuclei and Nucleolus, and other features.

All the images in these collections were shuffled so that, during the training process, the network would not see only specific categories of data, and each batch of images would contain different labels belonging to B-ALL and non-B-ALL categories. The size of the input image was changed to 300×300×3, but this method can be used on images with any size. The dataset was divided into training, validation, and test sets, according to the default hypothesis of [ALL challenge dataset of ISBI 2019](https://wiki.cancerimagingarchive.net/display/Public/C_NMC_2019+Dataset%3A+ALL+Challenge+dataset+of+ISBI+2019), whereby 10,661 of the data (ALL (cancer): 7272, normal: 3389, including 10% of the training dataset for validation) were allocated to the training set, and the remaining 1847 to the test set.

**2.3 Deep Learning Algorithms**

For medical image segmentation, numerous CNN-based methods are widely used benefitting from its powerful feature learning and representation ability. Of these methods, the convolutional Neural network (CNN) has demonstrated state-of-the-art performance in cells and organ segmentation problems^19–21^. CNN is a multi-layer network composed of overlapping convolutional layers (for feature extraction) and down-sampling layers (for feature processing). Figure 3 shows the structure of a typical convolutional neural network. CNN can automatically extract features from images, so it has become a research hotspot. CNN is a perceptron-based mode. CNNs can automatically extract features from images, and thus have become a research hot topic. CNNs are perceptron-based models^22,23^.

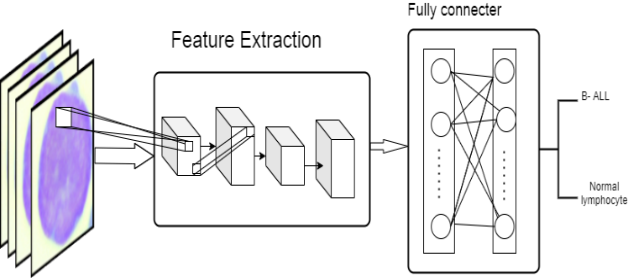
 Pre-trained models who have been trained in large image collections have achieved extraordinary results in their classic image issues and are therefore referred to as the State of the art. Many studies have used these top-notch models because their ability to extract image features has outperformed other models. Among the many publicly available deep learning pre-training models, Alexnet^24^, ResNet^25^(ResNet50, ResNet101 Inception-V3^25^, Inception-ResNet-V2^26^, SqueezeNet^27^, and MobileNet-V2^28^ Used and compared, because these network models show higher performance in terms of accuracy compared to any other networks with similar prediction times. Those well-known CNN models were pre-trained with the ImageNet database.

Figure 3: An ordinary convolutional neural network structure

$$\boldsymbol{Sensitivity=Recall=}\frac{\boldsymbol{True Positive}}{\boldsymbol{True Positive+False Negative}}\boldsymbol{Specificity=}\frac{\boldsymbol{True Negative}}{\boldsymbol{True Negative+False Posetive}}\boldsymbol{Accuracy=}\frac{\boldsymbol{True Negative+True Posetive}}{\boldsymbol{Totall Cases}}$$

3. **Results**

By implementation, seven models with learning data sets and then testing these models using a test dataset. It was concluded that different states of the art shown different performances. With respect to the similar features of the two data classes, the models did not reach an acceptable performance. The evaluation metric for the implemented network based on the confusion matrix is given in Table (2).

| *Table 2: Evaluation metrics of applied pre-Trained Models* | | | | |
| --- | --- | --- | --- | --- |
| GPU Time  (second) | Accuracy | Specificity | Sensitivity | Model |
| 4116 | 85.3% | 70.9% | 98.5% | DenseNet 121 |
| 8130 | 89.77% | 72.4% | 99% | Inception-V3 |
| 4966 | 78.6% | 69% | 88% | ***Alexnet*** |
| 5111 | 74% | 66% | 79% | ***ResNet-50*** |
| 10115 | 79% | 68% | 81% | ***ResNet-101*** |
| 24119 | 89.1% | 68.83% | 99.92% | Inception-ResNet-v2 |
| 5211 | 81% | 68.8% | 89% | ***SqueezeNet*** |
| 9232 | 94.91% | 85.65% | 99.84% | Xception |

Training these seven well-known models with training data sets and then validating them, four models achieved higher performance. These best four models (DenseNet121, Inception V3, Inception-ResNet-v2, and Xception) among eight models were selected by evaluating the performance of each model in terms of accuracy and calculation time.

**3.1 Ensemble deep learning model for classification B-ALL form B-lymphoblast (Normal)**

In this study, by examining the performance of the models used, the researchers sought a way to improve the results of these models. The majority-voting group was used as the final decision for this model. Present model scheme first calculates the total number of votes received by each base classifier, than the majority of votes is calculated by the classification of both class. Algorithm 1 presents the proposed model in detail. Let *L= {*DenseNet121, Inception V3, Inception-ResNet-v2 and Xception} be the set of pre-trained models. Four used selection model is fine tuned with the images from training datasets (*X; Y*); where *X* the set of *N* images, each of size, 300 × 300, and *Y* contain the images labels, *Y is a collection of two class including Lymphoblast B-ALL and Normal B-ALL*. After dividing the training dataset into batches size n=256.

***Algorithm 1:*** *Proposed Ensemble Algorithm*

Data: Lymphoblast Images(X,Y), X is the number of dataset images, Y={y$\in(Normal B-ALL, Lymphoblast B-ALL)$}

Result: Proposed model based Majority Voting technique that classifies class of lymphocyte images *x*$\in$ *X*

Applied Preprocessing

*Step 1: Resize images to dimension 300 × 300*

*Step 2: dataset image normalization , each image is mapped to [0,255] and then converted to the [0,1] for x*$\in$ *X*

*Step 3: Data set Edge detection and Tune Brightness and Contrast for x*$\in$ *X*

*Step 4: Data augmentation for every x*$\in$ *X(Rotation, filpping,cropping ,color transformation)*

Implement a set of pre-trained Model M={DenseNet121, Inception V3, Inception-Reset-v2,ResNet-50,ResNet101-Xception, SqueezeNet, Alexnet}

Models K={DenseNet121, Inception V3, Inception-Reset-v2,ResNet-50,ResNet101-Xception, SqueezeNet, Alexnet} with problem scale

**for each**	$k\in K$, α=0.0017, epochs=700

**for** i =1 to epochs

**for each** batch **do**

**for** calculation output and update the hyperparameter

**if** error is not improving change

using drop out, validation and feed to softmax

**end**

**end**

**end**

**end**

for each *x*$\in$ *xtest do*

*voting based majority performance for all models*

*end*

In this method, the number of base classifiers should always be odd. In the case of an equal vote, the mode function applies is applied.^36^ In this section, the proposed B-ALL disease detection and classification model is presented. The proposed model is displayed in Figure 4. It is an ensemble framework of four models.

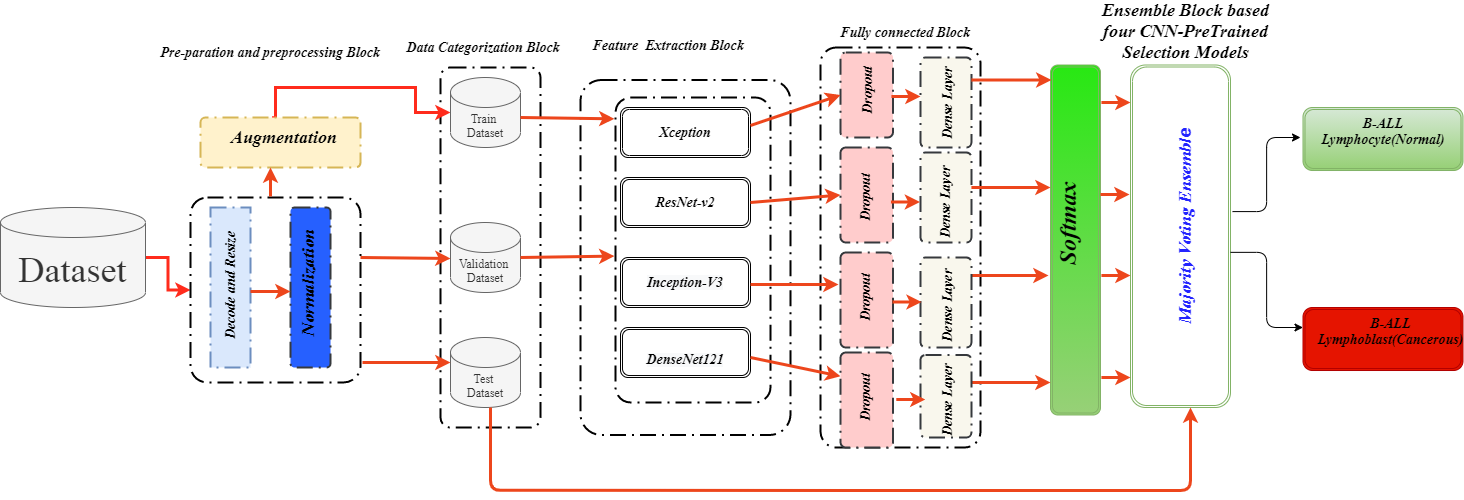

Figure 4: Proposed model for the classification of B-ALL lymphoblast from B-ALL lymphocytes

These four hybrid models are in parallel and combine module the output by the ensemble technique to improve the classification confidence and accuracy. Based on the confusion matrix in Figure 5, specificity and sensitivity are presented in Table 3 to assess the performance of the proposed ensemble model. The accuracy of the proposed model is 98.5% and the f1–score is 98.3%.The performance of the classification results of the proposed ensemble model and the previous state-of-the-art models is compared in Table (3). Evidently, the proposed ensemble model has achieved a promising performance that is superior to the previous models. The success of the proposed model with such a small data set is attributed to the use of class weights in the training process.

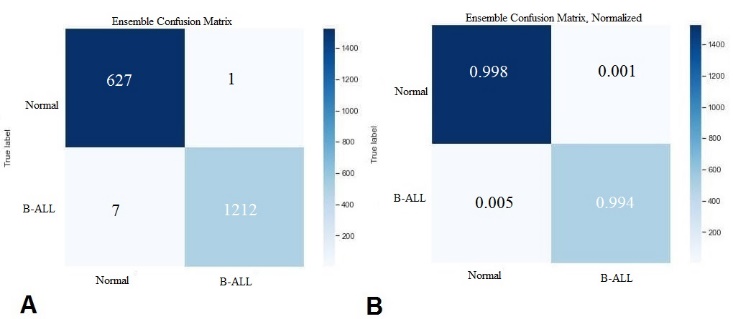

Figure 5:A. Confusion matrix, and B. Normalized confusion matrix of model performance

### 4. Discussion and Conclusion

| Table 3: Performance comparison of different models for ALL the detection | | | |
| --- | --- | --- | --- |
| Accuracy | Specificity | Sensitivity | Researcher, Year |
| 94.73 | 95.00 | 94.93 | Mohapatra S et al. (2013) |
| 89.72 | 95.08 | 80.43 | Singhal et al. (2014) |
| 96.06%. | 99.03 | 96.74% | Shafique S et al. (2018) |
| 98.1 | 77.85 | 88.5% | Yu et al. (2017) |
| 81.74 | Not Mentioned | 81.56 | Pansombut T et al.(2019) |
| 96.17 | 98.58 | 95.17 | Hosseinzadeh S. et al(2019) |
| **98.5%** | **96.7%** | **99.4** | **Proposed Model** |

The analysis of peripheral blood smear images plays a vital role in the diagnosis of various types of leukemia, anemia, and malaria. Unusual alterations in the color, shape, and size of blood cells indicate an abnormal condition. The results of this assessment, which is usually performed manually, depend on the skill and experience of the technician. Besides, manual microscopic examination is time-consuming and yields poor results^37–39^. The proposed method presented a framework for the automated classification of leucocyte cell nuclei from microscopic peripheral blood smear images. This study introduced a novel combination of image processing methodologies based on state-of-the-art CNNs. The ensemble framework based on majority voting for B-ALL detection using blood smear images achieved high accuracy in the classification of the two classes. Ensemble learning can significantly improve the generalization ability of the learning system, thereby enhancing the performance of the available methods.

**5. Acknowledgments**

Not applicable.

**Authors’ contributions**

Conceptualization:MG,AH.Programming of the frameworks:MG,AR,FA.Medical­ Supervisor:DB,HA. Writing,review and editing:MG,AH,FA.

**Funding**

This study was part of a PhD project conducted at Shahid Beheshti University of Medical Sciences, Tehran, Iran, was approved by Iran National Committee for Ethics in Biomedical Research with Approval ID IR.SBMU.RETECH.REC.1399.735.

**Availability of data and materials**

Data are available and public in<https://wiki.cancerimagingarchive.net/display/Public/C_NMC_2019+Dataset%3A+ALL+Challenge+dataset+of+ISBI+2019>

**Declarations**

**Ethics approval and consent to participate**

Not applicable.

**Consent for publication**

Not applicable.

**Competing interests**

The authors declare that they have no competing interest

8.	Dasariraju S, Huo M, Mccalla S. Detection and Classification of Immature Leukocytes for Diagnosis of Acute Myeloid Leukemia Using Random Forest Algorithm.

9.	Fathi E, Rezaee MJ, Tavakkoli-Moghaddam R, Alizadeh A, Montazer A. Design of an integrated model for diagnosis and classification of pediatric acute leukemia using machine learning. Proceedings of the Institution of Mechanical Engineers, Part H: Journal of Engineering in Medicine. doi:10.1177/0954411920938567

10.	Fan H, Zhang F, Xi L, Li Z, Liu G. LeukocyteMask : An automated localization and segmentation method for leukocyte in blood smear images using deep neural networks. doi:10.1002/jbio.201800488

11.	Gupta A, Gupta R. *Isbi 2019 C-Nmc Challenge: Classification in Cancer Cell Imaging*. Springer; 2020.

12.	Gehlot S, Gupta A, Gupta R. SDCT-AuxNetθ: DCT augmented stain deconvolutional CNN with auxiliary classifier for cancer diagnosis. *Med Image Anal*. 2020;61:101661. doi:https://doi.org/10.1016/j.media.2020.101661

13.	Goswami S, Mehta S, Sahrawat D, Gupta A, Gupta R. Heterogeneity Loss to Handle Intersubject and Intrasubject Variability in Cancer. *arXiv Prepr arXiv200303295*. Published online 2020.

14.	Clark K, Vendt B, Smith K, et al. The Cancer Imaging Archive (TCIA): maintaining and operating a public information repository. *J Digit Imaging*. 2013;26(6):1045-1057.

15.	Litjens G, Kooi T, Bejnordi BE, et al. A survey on deep learning in medical image analysis. *Med Image Anal*. 2017;42:60-88. doi:10.1016/j.media.2017.07.005

16.	Patro S, Sahu KK. Normalization: A preprocessing stage. *arXiv Prepr arXiv150306462*. Published online 2015.

17.	Frühwirth‐Schnatter S. Data augmentation and dynamic linear models. *J time Ser Anal*. 1994;15(2):183-202.

18.	Shorten C, Khoshgoftaar TM. A survey on Image Data Augmentation for Deep Learning. doi:10.1186/s40537-019-0197-0

19.	Ronneberger O, Fischer P, Brox T. U-net: Convolutional networks for biomedical image segmentation. In: *International Conference on Medical Image Computing and Computer-Assisted Intervention*. Springer; 2015:234-241.

20.	Lu L, Zheng Y, Carneiro G, Yang L. Deep learning and convolutional neural networks for medical image computing. *Adv Comput Vis Pattern Recognit*. Published online 2017.

27.	Iandola FN, Han S, Moskewicz MW, Ashraf K, Dally WJ, Keutzer K. SqueezeNet: AlexNet-level accuracy with 50x fewer parameters and< 0.5 MB model size. *arXiv Prepr arXiv160207360*. Published online 2016.

28.	Sandler M, Howard A, Zhu M, Zhmoginov A, Chen L-C. Mobilenetv2: Inverted residuals and linear bottlenecks. In: *Proceedings of the IEEE Conference on Computer Vision and Pattern Recognition*. ; 2018:4510-4520.

29.	Chung M, Bernheim A, Mei X, Zhang N, Radiology MH-, 2020 undefined. CT imaging features of 2019 novel coronavirus (2019-nCoV). *pubs.rsna.org*. 2020;295(1):202-207. doi:10.1148/radiol.2020200230

30.	Szegedy C, Vanhoucke V, Ioffe S, Shlens J. *Rethinking the Inception Architecture for Computer Vision*.; 2016.

31.	Szegedy C, Ioffe S, Vanhoucke V, Alemi A. Inception-v4, inception-resnet and the impact of residual connections on learning. In: *Proceedings of the AAAI Conference on Artificial Intelligence*. Vol 31. ; 2017.
